# Assessing Large Language Model Utility and Limitations in Diabetes Education: A Cross-Sectional Study of Patient Interactions and Specialist Evaluations

**DOI:** 10.1101/2025.06.24.25329401

**Authors:** Ghulam Mustafa, Joshua Ong, M. Zaman Shaikh, Saima Askari, Sarwat Anjum, Mohammad Idrees Adhi, Abdul Sami Memon, Muhammad Uzair Abdul Rauf, Arjumand Rizvi, Imran Iqbal, Shahla Basit, Muhammad Fahadullah Khan, Muhammad Qamar Masood

## Abstract

**Objectives:** To assess the value of an AI-powered conversational agent in supporting diabetes self-management among adults with diabetic retinopathy and limited educational backgrounds.

**Methods:** In this cross-sectional study, 51 adults with Type□II diabetes and diabetic retinopathy participated in moderated Q-and-A sessions with ChatGPT. Non-English-speaking and visually impaired participants interacted through trained human support. Each question– response pair was assigned to one of seven thematic categories and independently evaluated by endocrinologists and ophthalmologists using the 3C□+□2 framework (clarity, completeness, correctness, safety, recency). Inter-rater reliability was calculated with intraclass correlation coefficients (ICC) and Fleiss’□Kappa.

**Results:** The cohort generated 137 questions, and 98□% of the conversational agent’s answers were judged informative and empathetic. Endocrinologists awarded high mean scores for clarity (4.66/5) and completeness (4.52/5) but showed limited agreement (ICC□=□0.13 and□0.27). Ophthalmologists gave lower mean scores for clarity (3.09/5) and completeness (2.94/5) yet demonstrated stronger agreement (ICC□=□0.70 and□0.52). Reviewers detected occasional inaccuracies and hallucinations. Participants valued the agent for sensitive discussions but deferred to physicians for complex medical issues.

**Conclusions:** An AI conversational agent can help bridge communication gaps in diabetes care by providing accurate, easy-to-understand answers for individuals facing language, literacy, or vision-related barriers. Nonetheless, hallucinations and variable specialist ratings underscore the need for continuous physician oversight and iterative refinement of AI outputs.

**Practice implications:** Introducing conversational AI into resource-limited clinics could enhance patient education and engagement, provided that clinicians review and contextualise the advice to ensure safety, accuracy, and personalisation. Future development should prioritise reducing hallucinations and bolstering domain-specific reliability so the tool complements, rather than replaces, professional care.

## 1. Introduction

Diabetes mellitus affects millions globally, with projections showing an increase from 529 million in 2021 to 1.31 billion by 2050 (1, 2). Despite this rise, over 80% of people with diabetes live in low- and middle-income countries (LMICs), where fewer than 10% receive guideline-recommended care (2). Pakistan, home to the third largest diabetic population worldwide, faces severe challenges—including limited access to healthcare, medication costs, and a low doctor-to-patient ratio (3, 4).

For individuals with diabetic retinopathy and low literacy levels, these issues are magnified. Many struggle to manage their condition due to language barriers and limited access to understandable health education.

Artificial intelligence (AI), especially large language models (LLMs) like ChatGPT, has emerged as a potential tool to address this gap (5, 6). By providing accessible and empathetic responses to health-related queries, AI tools could improve patient understanding and engagement (7, 8). While past research has explored LLMs in lab test interpretation (9), and rare disease queries (10), diabetes-focused chatbot studies often remain limited to dietary advice (11, 12), overlooking other key management areas like emotional support, comorbidities, and medication education.

Furthermore, many evaluations rely on simulated data (13). There’s limited understanding of how real users, especially those with limited education, engage with AI—and how physicians perceive the accuracy and safety of these interactions (14, 15).

We hypothesize that AI-generated responses, when translated and used under physician oversight, can support diabetes self-management in underserved populations. This includes those with diabetic retinopathy who face significant literacy and language challenges.

To test this, we developed the 3C□+□2 framework—evaluating chatbot responses based on clarity, completeness, correctness, and two additional pillars: safety and recency (alignment with current medical knowledge). This framework aims to set a standard for analyzing medical chatbot content.

This study explores the educational value of ChatGPT beyond dietary advice, focusing on real interactions from individuals with diabetic retinopathy in Karachi, Pakistan. By evaluating ChatGPT’s performance through a physician’s lens, we provide insights on integrating AI into diabetes care—especially in settings with limited resources.

## 2. Method

This cross-sectional study enrolled adults with Type II diabetes attending medical consultations at hospitals in Karachi, Pakistan. Ethical approval was granted by the Institutional Review Board of the Pakistan Council of Scientific and Industrial Research (PCSIR) Laboratories Complex, Karachi, Pakistan (Ref. IES/HS-02; January 2, 2023); by the Research Ethics Committee of Al-Ibrahim Eye Hospital, Karachi, Pakistan (Ref. REC/IPIO/2023/070; January 5, 2023); and by the Ethics Committee of Fatimiyah Hospital, Karachi, Pakistan (Ref. HR/FH-TZ-090824; April 3, 2023). The study was registered at ClinicalTrials.gov (Identifier: NCT05883072). All participants provided written informed consent prior to enrollment.

Following ethical approval, we enrolled people with diabetes between January 17, 2023, and April 26, 2024. Each participant was fully briefed on the study details and provided written informed consent prior to participation. Only those who provided written informed consent were included in the study. (Figure 1)

**Figure 1.**
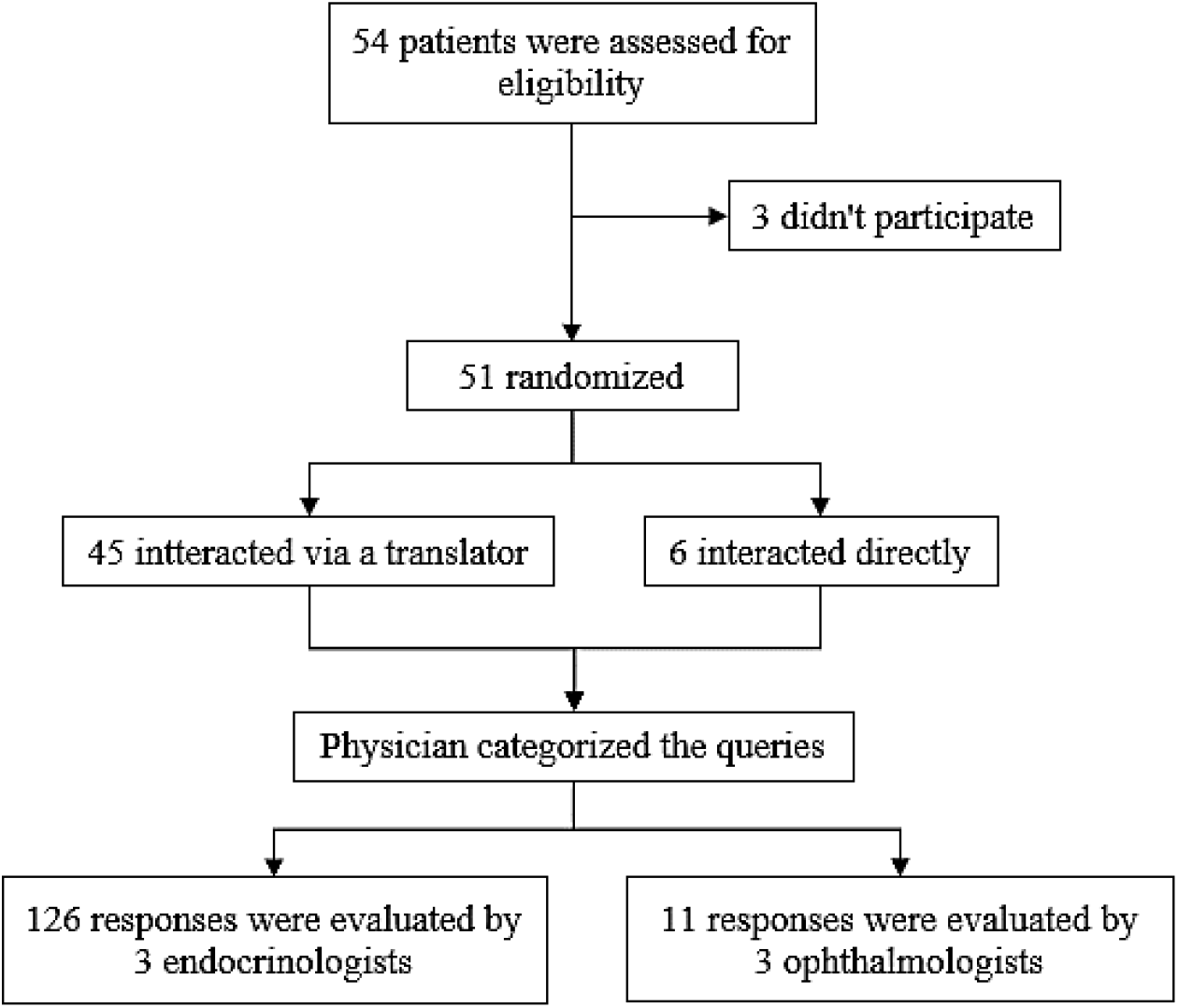
Consort Diagram

We collected basic demographic and background information from each participant, including age, education, healthcare expenditure, and family history of diabetes. Participants interacted with ChatGPT via the Chrome browser on a Huawei AL00 Android mobile phone. For participants proficient in English, the interaction was conducted directly. For individuals not proficient in English, the Principal Investigator acted as an interpreter, interpreting the participants’ queries into English and relaying ChatGPT’s responses back to them in Urdu. Each interaction lasted approximately 15 to 45 minutes, depending on the participant’s interaction method.

Some interactions began with prompts such as “I am a diabetic patient” or “I am a type 2 diabetic patient” to ensure responses were tailored to diabetes-specific queries. All participants had a confirmed diagnosis of diabetes and were following various treatment guidelines. They asked questions related to blood sugar management, prevention and care, diet and nutrition, diabetic complications and symptoms, medications and treatment, and lifestyle and daily management. The number of questions per participant ranged from 1 to 8, depending on the time available, level of interest, and satisfaction with the information provided by ChatGPT.

After the interaction, participants were asked to provide feedback on their experience with ChatGPT. Specifically, they rated it on being informative, detailed, and empathetic. Additionally, they were asked whether they had used ChatGPT before and if they would prefer to use it for medical guidance in the future. These responses were collected as binary inputs.

All interactions took place either in the presence of a medical consultant or just before their consultation. This was done to mitigate the potential impact of misinformation (i.e., hallucinations) generated by ChatGPT, ensuring that no harmful knowledge was imparted to the individuals.

Each interaction was recorded for subsequent evaluation by medical specialists. A physician first reviewed the recorded chats and categorized them into two groups for further analysis. Chats related to diabetic retinopathy were sent to an ophthalmologist, while all other chats were sent to an endocrinologist.

Each group of medical specialists (ophthalmologists and endocrinologists) consisted of three professionals with between 3 and 20 years of medical experience. These specialists were tasked with analyzing the responses generated by ChatGPT using a 3C framework: Clarity, Completeness, and Correctness. Clarity was defined as the ease with which the person with diabetes could understand the information provided. Completeness referred to whether all necessary information was communicated to address the individual’s inquiry. Correctness was assessed by verifying that the information was accurate and did not contain any errors. The specialists were also asked to determine whether the information was safe and up-to-date.

Specialists rated clarity and completeness on a 1-5 scale, with 1 being “Very Poor” and 5 being “Very Good.” Correctness, safety, and up-to-dateness were evaluated as binary (Yes/No) variables. Specialists were encouraged to provide comments if they disagreed with ChatGPT’s responses, identifying areas where information was incomplete or inaccurate. The final scores for clarity and completeness were calculated by averaging the ratings, while correctness, safety, and up-to-dateness were assessed using a parity check.

In cases where specialists did not provide a full score—such as marking a response as “Poor” without offering specific suggestions for improvement—the chats were returned to the specialists for clarification. The specialists were requested either to adjust their rating or provide a rationale for their initial assessment.

To evaluate the reliability of the ratings provided by the specialists, we calculated the Intraclass Correlation Coefficient (ICC) using a two-way random-effects model with average measures. The ICC in this model evaluates how strongly units in the same group resemble each other when both subjects and raters are treated as random factors. We used the ICC to assess the consistency of ratings within the same group for the dimensions of Clarity and Completeness.

ICC was used to evaluate the consistency of ratings among multiple specialists. We used open-source python library to calculate the ICC.

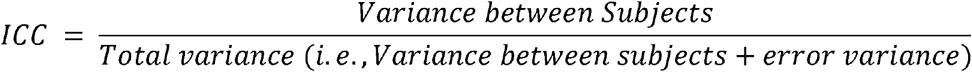

For the evaluation of Correct, Safe, and Up-to-date ratings, which were binary in nature, we employed Fleiss’ Kappa. This method measures the degree of agreement among two or more raters, while adjusting for chance agreement. Python code was written to compute both ICC and Fleiss’ Kappa.

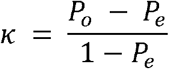

In order to further investigate areas with low ICC among the endocrinologists, we extracted 10 chats in which all three endocrinologists agreed and 11 chats where at least one of them disagreed. These chats were reviewed by another endocrinologist with 10 years of experience. The new specialist’s analysis revealed disagreements with both the previously unanimous ratings and those where disagreement had already been noted, highlighting the subjective nature of these evaluations.

## 3. Results

### 3.1 Participants Queries

Initially, 54 participants consented to participate in the study; however, three did not submit any queries to ChatGPT and were subsequently excluded. A total of 137 queries were received from the remaining 51 individuals with diabetes, averaging over two questions per individual. The number of queries per individual ranged from one to eight. (Figure 2) To gain deeper insights into the participants’ areas of interest, these queries were categorized into seven distinct groups (Figure 3).

**Figure 2.**
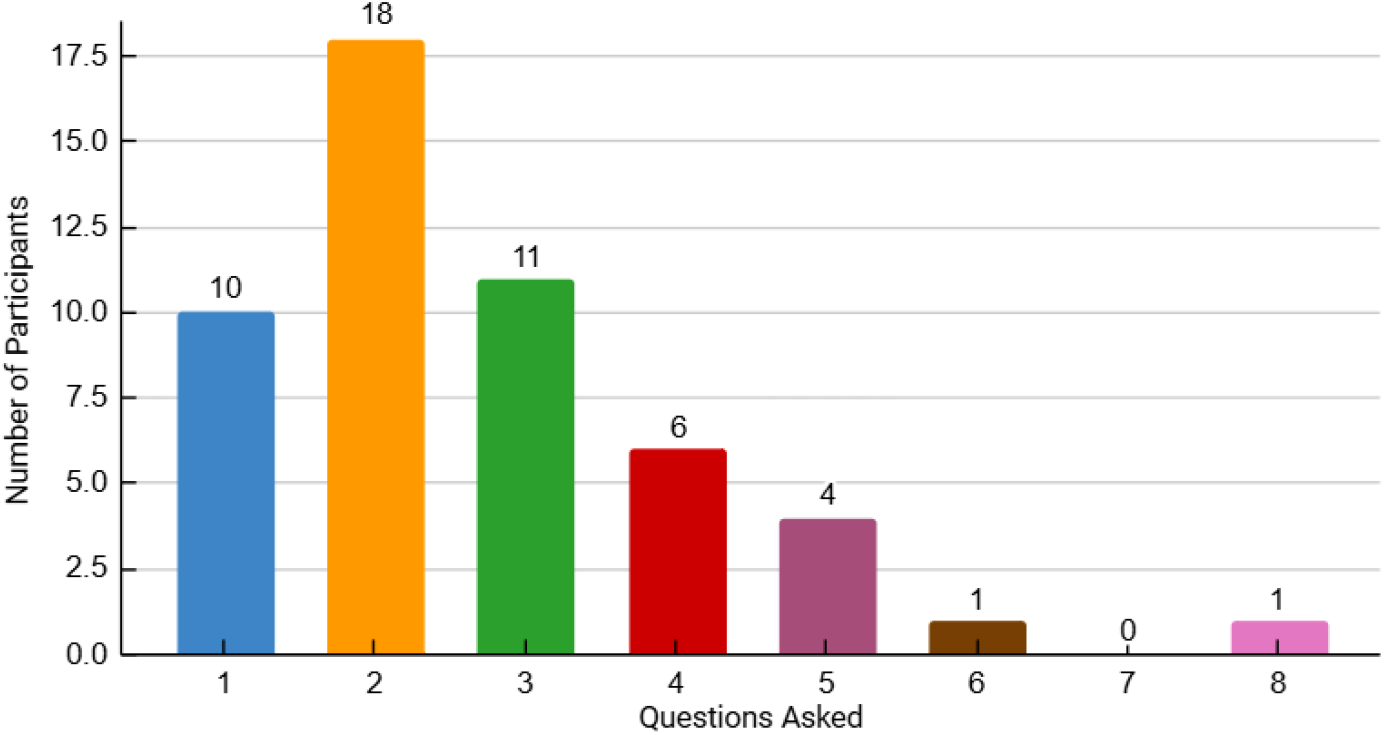
Distribution of Participants by Number of Queries

**Figure 3.**
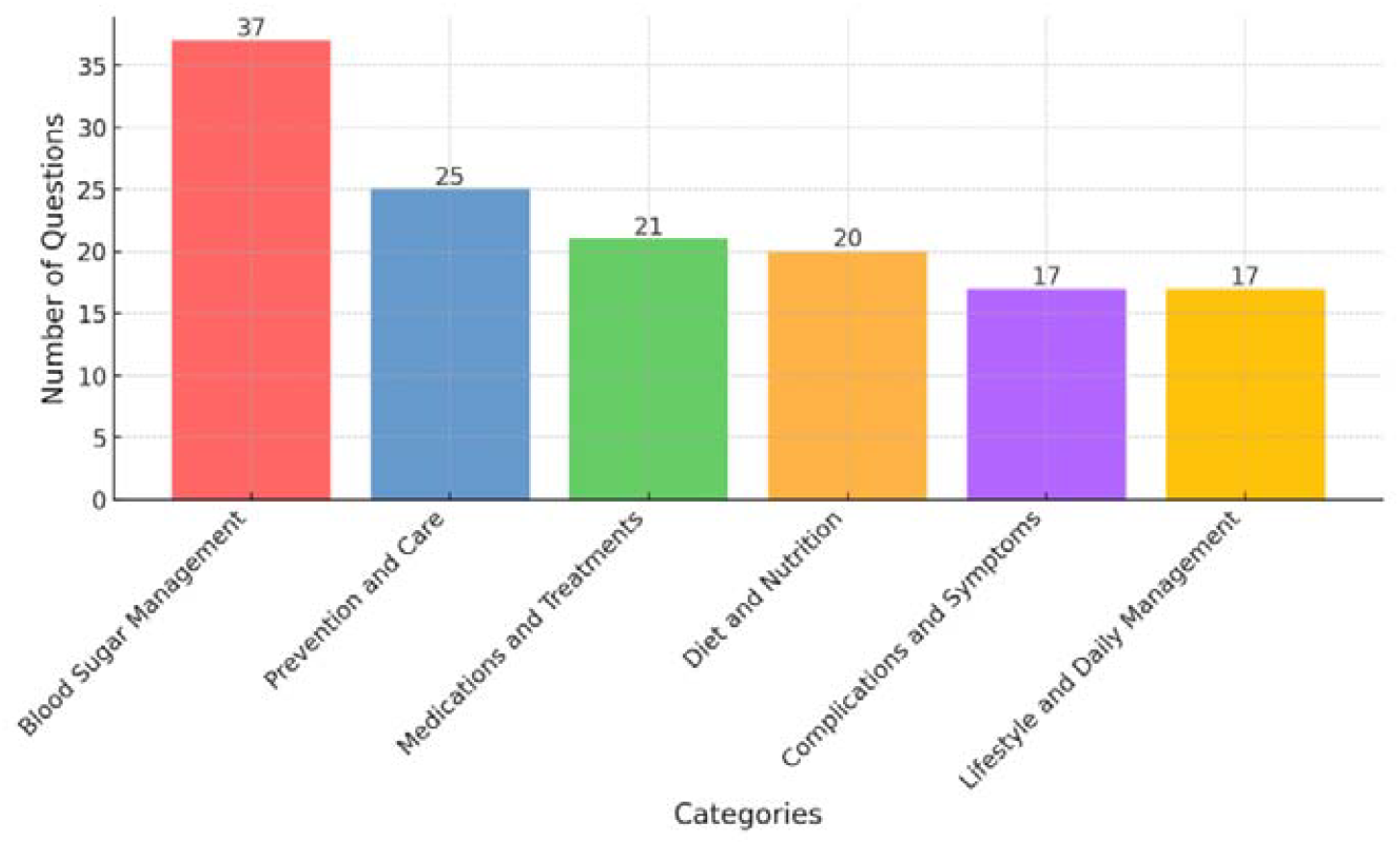
Categories of Questions Asked by Participants

#### 3.1.1 Participants Demographics

Among the participants, 53.97% were male, and 46.03% were female. Their ages ranged from 19 to 70 years, with educational backgrounds spanning from no formal education to a master’s degree (Figure 4). The duration of diabetes among participants varied from 3 months to 35 years. A family history of diabetes was reported by 57.4% of the participants, with 28.1% having both parents diagnosed with the condition (Table 1).

**Table 1:**
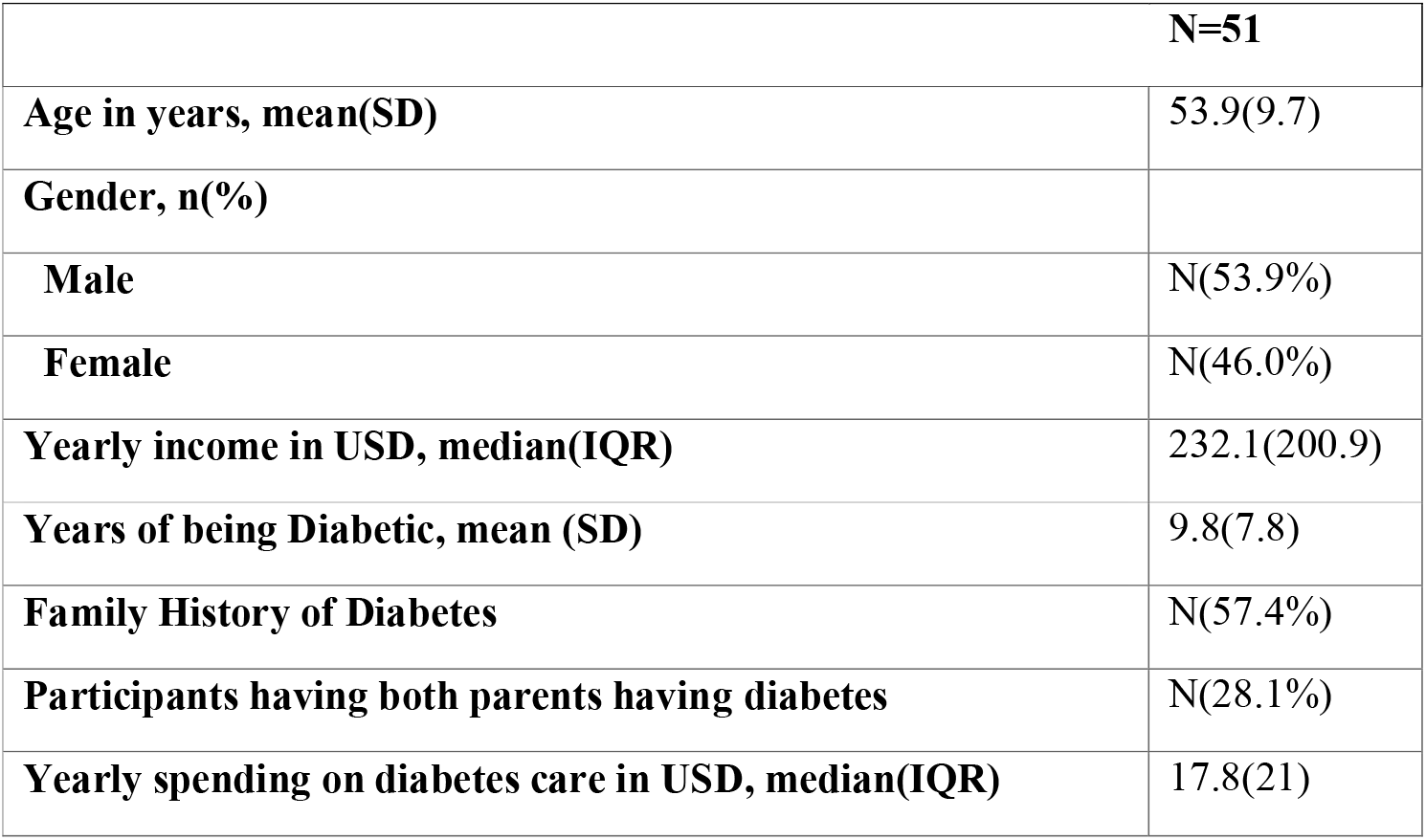
Baseline Characteristic of study participants.

**Figure 4.**
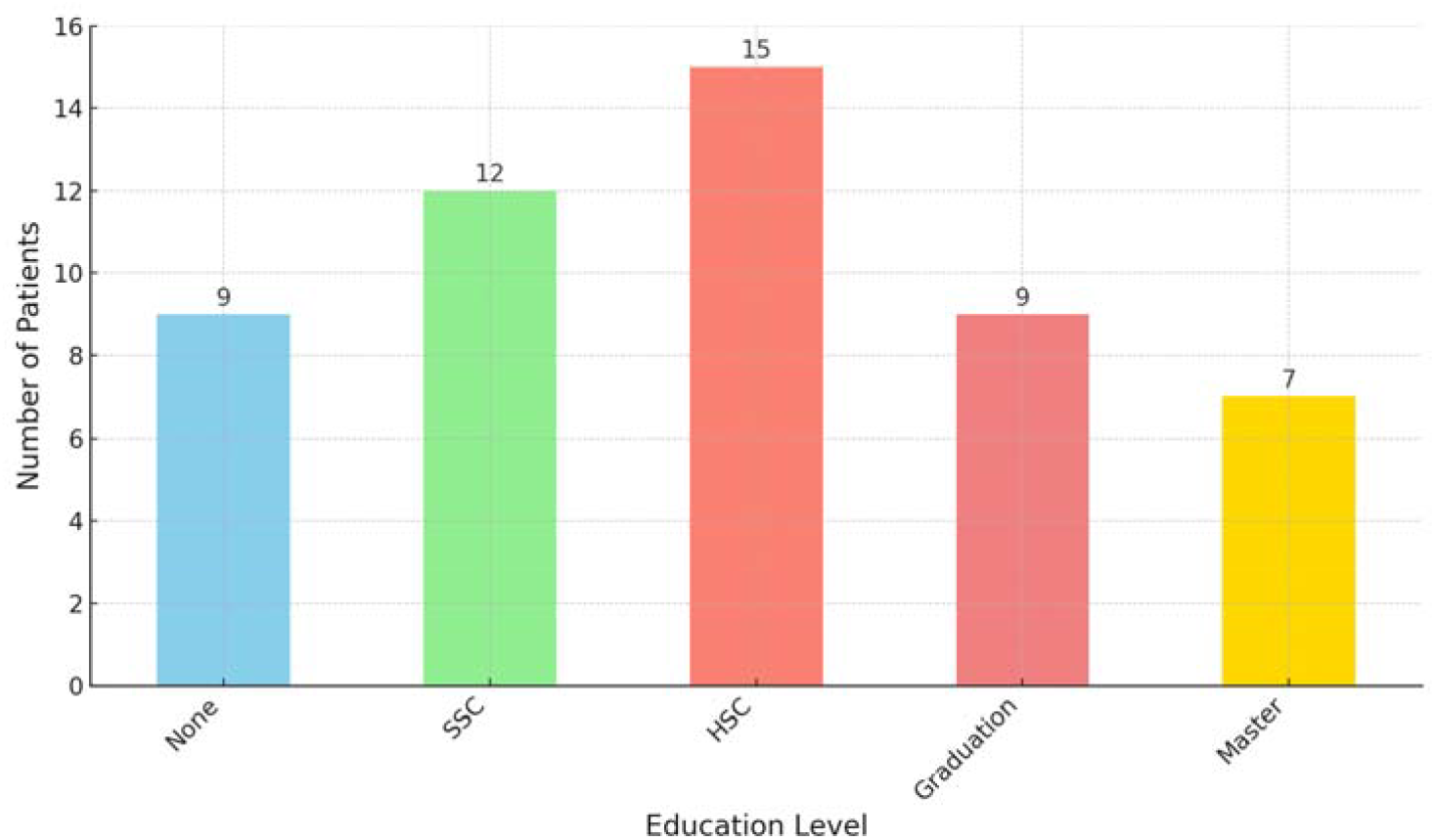
Distribution of Education Levels Among Study Participants.

None (no formal education), SSC (secondary school certificate—10 years of education), HSC (high school certificate—12 years of education), Graduation (college degree—14-16 years of education), Masters (16-18 years of education).

#### 3.1.2 Financial Status

We assessed the financial situation of participants, finding that 43.4% had no source of personal income and were dependent on family members, particularly in the case of housewives. The yearly earnings disclosed by individuals ranged from $171 to $10,800. Yearly spending on diabetes-related care ranged from $0 to $4,300 (Table 1).

### 3.2 Participants Feedback

Only 3.9% of individuals reported having prior experience with ChatGPT; however, all expressed interest in utilizing it for future consultations. These people universally found ChatGPT’s responses to be empathetic, with 98% considering the information both informative and detailed. The single critical comment from a participant was due to receiving advice from ChatGPT that differed from their physician’s, which led them to perceive the information as less informative and detailed.

### 3.3 Specialist Evaluation

A total of 126 ChatGPT responses (s1) were independently evaluated by three endocrinologists (s2, s3, s4), while 11 were assessed by three ophthalmologists (s5, s6, s7). Responses were rated for clarity and completeness (on a 5-point scale) and correctness, safety, and up-to-date (as yes/no).

#### 3.3.1 Endocrinologist Ratings

The average clarity score given by the endocrinologists was 4.66 (SD = 0.45), with a minimum of 3.33 and a maximum of 5. Completeness received an average score of 4.52 (SD = 0.56), with a range from 3 to 5. The intraclass correlation coefficient (ICC) for clarity was 0.13, while completeness had an ICC of 0.27 (Table 2). Fleiss’ Kappa values for Correct, safe and latest were 0.05, 0.09, and −0.05 respectively. The highest agreement among the three endocrinologists revealed that 98.4% of responses were correct, 100% were safe, and 100% were up-to-date. The percentage of agreement among the endocrinologists was 98.41%, 99.2%, and 100% for completeness, safety, and currency, respectively (Table 3).

**Table 2:**
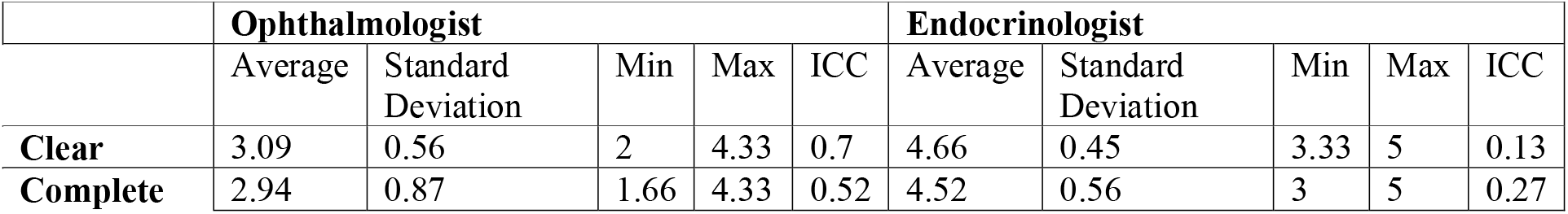
Evaluation Metrics Summary: Clarity and Completeness Ratings.

**Table 3:** Evaluation of Response Accuracy, Safety, and Latest Ratings.

|  | Ophthalmologist |  | Endocrinologist |  |
| --- | --- | --- | --- | --- |
|  | Percentage | Fleiss Kappa | Percentage | Fleiss Kappa |
| Correct | 54.54 | 0.38 | 98.41 | 0.05 |
| Safe | 81.81 | 0.67 | 99.2 | 0.09 |
| Latest | 63.63 | 0.14 | 100 | -0.05 |

#### 3.3.2 Ophthalmologist Ratings

The average clarity score from the ophthalmologists was 3.09 (SD = 0.56) with a minimum as 2 and maximum as 4.33, and completeness scored 2.94 (SD = 0.87) with a minimum of 1.66 and maximum of 4.33. The ICC for clarity was 0.7, indicating good inter-rater reliability, while completeness had an ICC of 0.52 (Table 2). The percentage of agreement among the ophthalmologists was 54.55% for correctness, 81.82% for safety, and 63.63%% for up-to-date. The corresponding Fleiss’ Kappa values were 0.36, 0.82, and 0.13, respectively. Agreement between two graders showed that 70% of the chats were deemed correct, 80% safe, and 80% up-to-date (Table 3).

## 4. Discussion

This study is the first to evaluate ChatGPT 3.5’s ability to address medical queries from people with diabetes. We analyzed chatbot-patient interactions, response quality, and expert assessments from an endocrinologist and an ophthalmologist. Additionally, we created a novel dataset that offers valuable insights for both AI and medical research.

Our findings highlight ChatGPT’s potential in diabetes education, particularly for people with limited literacy and English proficiency. Despite only 3.7% of participants being familiar with ChatGPT beforehand, they found its responses detailed and empathetic. The model’s ability to handle multilingual scenarios demonstrated its versatility in offering health guidance across various languages.

However, participants preferred physician advice, especially for complex queries like the reversibility of diabetic conditions, emphasizing AI’s current limitations in handling nuanced health questions. Internet connectivity issues also hindered interactions, pointing to the need for an offline version to improve reliability.

Participants felt more comfortable discussing sensitive issues like sexual dysfunction with ChatGPT, particularly when gender differences made in-person consultations awkward. This suggests that chatbots could help overcome privacy barriers in healthcare.

A major concern was the occurrence of hallucinations, where ChatGPT generated incorrect or unsafe advice. While participants found these responses satisfactory, experts flagged them as potentially harmful, highlighting the need for improved reliability in AI-generated medical information.

GPT-4, though generally safe, often preferred newer, more expensive drugs like GLP-1 receptor agonists over metformin, raising equity and cost concerns (16). This emphasizes the need for careful prompt design and refinement to avoid biases in medical recommendations.

We also observed variability in specialist evaluations of ChatGPT’s responses, reflecting differences in medical interpretations and expectations, making it difficult to standardize AI outputs in clinical practice.

Human-in-the-Loop (HITL) systems offer a promising solution, where AI suggestions are reviewed by clinicians to ensure relevance and safety (17). This collaborative approach can combine AI efficiency with human judgment.

Data security was a critical concern, following an incident involving the exposure of sensitive user information. This highlights the importance of robust safeguards for AI technologies in clinical settings.

While AI can support patient education, it cannot replace physicians, particularly due to limitations in accountability, personalization, and contextual understanding. Open-source, well-regulated LLMs will be essential for safe and reliable AI-driven medical advice.

Current chatbots lack the ability to incorporate patient histories, leading to less personalized health advice. Future versions should focus on gathering this information to provide tailored recommendations.

The dataset generated in this study is an invaluable resource for future research, offering a foundation for evaluating emerging chatbots and improving AI integration into patient care. It will also facilitate comparisons with future iterations of ChatGPT and help advance AI in medical education.

Future studies may also consider incorporating HITL frameworks and prompt engineering strategies, as seen in recent GPT-4 research, to improve the consistency and safety of AI responses while maintaining adaptability to evolving clinical guidelines (17).

## 5. Conclusion

This study underscores the potential of ChatGPT as a supplementary tool for educating people with diabetes, particularly those with limited literacy and language proficiency. While the chatbot was effective in addressing basic queries and fostering patient engagement, its limitations—such as hallucinations, lack of contextual understanding, and variability in specialist assessments—highlight the need for cautious implementation.

ChatGPT’s utility was further constrained by internet connectivity issues, particularly in resource-limited settings, and concerns regarding data security. Future developments in AI-driven medical tools must prioritize reliability, personalization, and robust data protection. This research provides a valuable dataset for advancing AI in healthcare and sets the stage for further exploration of integrating AI with traditional medical practices to improve patient outcomes.

## Data Availability

All data produced in the present work are contained in the manuscript

## Acknowledgment

**Personal Thanks**. The author would like to specially acknowledge Fatimiyah Hospital for their invaluable support in this study. Sincere gratitude is also extended to the following individuals for their contributions to achieving this result: Dr. Ali Asghar, Consultant Endocrinologist and President-Elect of the Pakistan Endocrine Society; Syeda Maria Azfar, Biomedical Engineer Intern at PCSIR; Dr. Murtaza Ali Gowa, Department of Pediatric Medicine, National Institute of Child Health, Karachi, Pakistan; and Asad Ali, CEO of Husaini Blood Bank.

## References

1. Ling W, Huang Y, Huang Y-M, Fan R-R, Sui Y, Zhao H-L. Global trend of diabetes mortality attributed to vascular complications, 2000–2016. Cardiovascular Diabetology 2020;19 10.1186/s12933-020-01159-5

2. Sheng B, Pushpanathan K, Guan Z, Lim QH, Lim ZW, Yew SME, Goh JHL, Bee YM, Sabanayagam C, Sevdalis N, Lim CC, Lim CT, Shaw J, Jia W, Ekinci EI, Simó R, Lim L-L, Li H, Tham Y-C. Artificial intelligence for diabetes care: current and future prospects. The Lancet Diabetes & Endocrinology 2024;12:569–595 10.1016/S2213-8587(24)00154-2

3. Sun H, Saeedi P, Karuranga S, Pinkepank M, Ogurtsova K, Duncan BB, Stein C, Basit A, Chan JCN, Mbanya JC, Pavkov ME, Ramachandaran A, Wild SH, James S, Herman WH, Zhang P, Bommer C, Kuo S, Boyko EJ, Magliano DJ. IDF Diabetes Atlas: Global, regional and country-level diabetes prevalence estimates for 2021 and projections for 2045. Diabetes Res Clin Pract 2022;183:109119 10.1016/j.diabres.2023.110945

4. Muhammad Q, Eiman H, Fazal F, Ibrahim M, Gondal MF. Healthcare in Pakistan: Navigating Challenges and Building a Brighter Future. Cureus 2023;15:e40218 10.7759/cureus.40218

5. Haug CJ, Drazen JM. Artificial Intelligence and Machine Learning in Clinical Medicine, 2023. N Engl J Med 2023;388:1201–1208 10.1016/j.ijmedinf.2025.105889

6. Loh, E. (2023). ChatGPT and generative AI chatbots: challenges and opportunities for science, medicine and medical leaders. BMJ leader, leader-2023. 10.1136/leader-2023-000797

7. Sharma S, Pajai S, Prasad R, Wanjari MB, Munjewar PK, Sharma R, Pathade A. A Critical Review of ChatGPT as a Potential Substitute for Diabetes Educators. Cureus 2023;15:e38380 10.7759/cureus.38380

8. Tian S, Jin Q, Yeganova L, Lai PT, Zhu Q, Chen X, Yang Y, Chen Q, Kim W, Comeau DC, Islamaj R, Kapoor A, Gao X, Lu Z. Opportunities and challenges for ChatGPT and large language models in biomedicine and health. Brief Bioinform 2023;25 10.1093/bib/bbad493

9. Cadamuro J, Cabitza F, Debeljak Z, De Bruyne S, Frans G, Perez SM, Ozdemir H, Tolios A, Carobene A, Padoan A. Potentials and pitfalls of ChatGPT and natural-language artificial intelligence models for the understanding of laboratory medicine test results. An assessment by the European Federation of Clinical Chemistry and Laboratory Medicine (EFLM) Working Group on Artificial Intelligence (WG-AI). Clin Chem Lab Med 2023;61:1158–1166 10.1515/cclm-2023-0355

10. Reese JT, Chimirri L, Danis D, Caufield JH, Wissink K, Casiraghi E, Valentini G, Haendel MA, Mungall CJ, Robinson PN. Evaluation of the Diagnostic Accuracy of GPT-4 in Five Thousand Rare Disease Cases. medRxiv 2024; 10.1101/2024.07.22.24310816

11. Sun H, Zhang K, Lan W, Gu Q, Jiang G, Yang X, Qin W, Han D. An AI Dietitian for Type 2 Diabetes Mellitus Management Based on Large Language and Image Recognition Models: Preclinical Concept Validation Study. J Med Internet Res 2023;25:e51300 10.2196/51300

12. Papastratis I, Konstantinidis D, Daras P, Dimitropoulos K. AI nutrition recommendation using a deep generative model and ChatGPT. Sci Rep 2024;14:14620 10.1038/s41598-024-65438-x

13. Bedi S, Liu Y, Orr-Ewing L, Dash D, Koyejo S, Callahan A, Fries JA, Wornow M, Swaminathan A, Lehmann LS, Hong HJ, Kashyap M, Chaurasia AR, Shah NR, Singh K, Tazbaz T, Milstein A, Pfeffer MA, Shah NH. Testing and Evaluation of Health Care Applications of Large Language Models: A Systematic Review. JAMA 2024; 10.1001/jama.2024.21700

14. Sng GGR, Tung JYM, Lim DYZ, Bee YM. Potential and Pitfalls of ChatGPT and Natural-Language Artificial Intelligence Models for Diabetes Education. Diabetes Care 2023;46:e103–e105 10.2337/dc23-0197

15. Hager P, Jungmann F, Holland R, Bhagat K, Hubrecht I, Knauer M, Vielhauer J, Makowski M, Braren R, Kaissis G, Rueckert D. Evaluation and mitigation of the limitations of large language models in clinical decision-making. Nat Med 2024;30:2613–2622 10.1038/s41591-024-03097-1

16. Flory JH, Ancker JS, Kim SYH, Kuperman G, Petrov A, Vickers A. Large language model GPT-4 compared to endocrinologist responses on initial choice of glucose-lowering medication under conditions of clinical uncertainty. Diabetes Care 2025;48:185–192. 10.2337/dc24-1067

17. Pavon JM, Schlientz D, Maciejewski ML, Economou-Zavlanos N, Lee RH. Large language models in diabetes management: the need for human and artificial intelligence collaboration. Diabetes Care 2025;48:182–184. 10.2337/dci24-0079

